# The comparison of handpiece anti-surge system and main body in cataract surgery

**DOI:** 10.1101/2023.10.18.23297207

**Authors:** Hyo Won Kim, Inkee Kim, Hyun Seung Kim, Eun Chul Kim

## Abstract

**Background/aims:** To compare the efficacy of the Centrion handpiece anti-surge system (Active Sentry®) and main body according to the grade of nucleosclerosis.

**Methods:** 600 eyes of 415 patients who underwent cataract surgery was retrospectively enrolled. Intraoperative parameters included phacoemulsification time (seconds), cumulative dissipated energy (CDE), and balanced salt solution (BSS) use (ml). Clinical measurements were made preoperatively and at one day, one month, two, and three months postoperatively, including the best corrected visual acuity (BCVA), and the corneal endothelial cell (CEC) count in the handpiece group (N=300) and main body group (N=300). Within the anti-surge group, the number of anti-surge system activated was collected.

**Results:** Anti-surge (times), phacoemulsification time, CDE, and BSS use significantly increased with increasing nucleosclerosis grades and in zonule weakness, poor mydriasis, and pseudoexfoliation syndrome in handpiece group, respectively (p < 0.05). Phacoemulsification time, CDE, and BSS use of handpiece group were significantly lower than those of main body group (p < 0.05). Phacoemulsification time, CDE, and CEC loss of handpiece group were significantly lower than those of main body group in nucleosclerosis grade 5 and 6 (p < 0.05). BSS uses of handpiece group were significantly lower than those of main body group in all nucleosclerosis grade (p < 0.05).

**Conclusion:** Because of the fast-reacting anti-surge, Intraoperative parameters were saved in handpiece anti-surge system rather than in main body. Therefore, the anti-surge system in the Centrion handpiece helps the surgeon perform safer cataract surgery in higher nucleosclerosis grade, zonule weakness, poor mydriasis, and pseudoexfoliation syndrome.

**Synopsis:** Anti-surge system in the Centrion handpiece helps the surgeon perform safer cataract surgery in higher nucleosclerosis grade, zonule weakness, poor mydriasis, and pseudoexfoliation syndrome.

**WHAT IS ALREADY KNOWN ON THIS TOPIC:** Active Sentry handpiece was reported to be as safe and efficacious as the Ozil handpiece, with the benefit of operating at lower intraocular pressure levels

**WHAT THIS STUDY ADDS:** Anti-surge system in the Centrion handpiece helps the surgeon perform safer cataract surgery in higher nucleosclerosis grade, zonule weakness, poor mydriasis, and pseudoexfoliation syndrome.

**HOW THIS STUDY MIGHT AFFECT RESEARCH, PRACTICE OR POLICY:** With anti-surge system in the Centrion handpiece, phacoemulsification can be performed effectively and safely in patients with complicated cataract.

## Introduction

A stable anterior chamber is an essential factor for successful cataract surgery.^1^ Phacoemulsification systems, fluidics, and ultrasound tips have been developed for safer surgery to keep the anterior chamber stable.^2^ Anterior chamber stability is controlled by balances in inflow and outflow, which are affected by postocclusion surge, positive pressure, compliance of the aspiration tube, phacoemulsification tips and sleeves, and the phacoemulsification machine’s responsiveness of the sensor.^3^ In the conventional gravity-based fluidic system, the flow rate of the irrigation fluid depends on adjusting the height of the bottle.^4^ But the active fluidics system compresses or decompresses the balanced salt solution fluid bag with two metal plates and adjusts the perfusion flow in time to maintain intraocular pressure (IOP) to compensate for flow losses.^3 5^

An occlusion can occur when the tip of a phacoemulsification probe is obstructed by lens fragments, iris tissue, or ophthalmic viscoelastic device material, leading to vacuum rise inside the aspiration and the collapse of the tubing.^6^ An occlusion break can cause an abrupt aspiration of lens materials from the phacoemulsification tip and a subsequent rapid inflow of fluid from the anterior chamber.^7^ The fluid surge can cause several complications, such as anterior chamber collapse, posterior capsule rupture, and vitreous loss.^8 9^ The fluid surge is prevented with the Active Sentry system of the Centurion Vision System (Alcon Laboratories, Inc.) with the venting mechanism. In earlier versions of the Active Sentry system, pressure alterations at the tip of the phacoemulsification probe traveled to a sensor near the cassette in the main body of the phacoemulsification machine and reduced IOP fluctuations.^10^ An Active Sentry handpiece containing an integrated pressure sensor inside the handpiece was developed to respond faster and more sensitively to changes in IOP by eliminating the delay in fluid adjustment compared to the earlier version.^11^

There were several studies to compare the Active Sentry handpiece and the Ozil handpiece.^10 12 13^ Active Sentry handpiece was reported to be as safe and efficacious as the Ozil handpiece, with the benefit of operating at lower intraocular pressure levels.^10 12^

To the best of our knowledge, no study has compared the effectiveness of the Centrion handpiece anti-surge system and main body based on the degree of nucleosclerosis and challenging conditions such pseudoexfoliation syndrome, zonule weakness, and suboptimal mydriasis.

Thus, the objective of this study was to compare the efficacy of the Centrion handpiece anti-surge system and main body, and evaluate the frequency of the anti-surge system in the Centrion handpiece in cataract surgery according to the grade of nucleosclerosis.

## Methods

We performed a retrospective chart review and data analysis in this study. This study was conducted in compliance with Institutional Review Board regulations and the Declaration of Helsinki. The Institutional Review Board (IRB)/Ethics Committee of Bucheon St. Mary Hospital approved this study protocol.

### Patients’ examination

A total of 600 eyes from 416 patients who underwent phacoemulsification and intraocular lens implantation at Bucheon St. Mary Hospital from April 2019 to March 2023 were enrolled. All patients were divided with the handpiece group (N=300) and the main body group (N=300). Their demographic and perioperative data were recorded after a complete ophthalmological examination. Cataract nucleosclerosis grading was done by the Lens Opacities Classification System III. Uncorrected and corrected visual acuities were expressed as logMAR. Manifest refraction, biometry, and keratometry with the IOLMaster partial coherence interferometry device (Carl Zeiss Meditec AG), corneal topography (Pentacam®, Oculus, Germany), slit lamp examination, and dilated funduscopy were examined at the preoperative period and postoperative 1 day, 1 week, 1 month, 2 month, and 3 months. Within the handpiece group, the number of anti-surge system activated was collected.

### Operative Procedures

A single skilled surgeon (E. C. K) operated on all patients under topical anesthesia using a CENTURION® (Alcon Laboratories, Inc., Fort Worth, TX, USA). A single skilled surgeon (E. C. K) operated all patients under topical anesthesia using a CENTURION® (Alcon Laboratories, Inc., Fort Worth, TX, USA). The corneal steep axis and 6.0mm ring were marked with Gentian violet. Surgery was performed through a clear corneal incision at the steep astigmatic axis. After topical ocular anesthesia was applied, a 2.75-mm clear corneal incision was made using a 2.75-mm double-blade keratome (Alcon). Surgically induced astigmatism was set at 0.5 diopters. Sodium hyaluronate 1.0% (Hyal Plus, LG Life Science, Seoul, Korea) was used to reform and stabilize the anterior chamber. A continuous curvilinear capsulotomy was made according to a 6.0mm corneal marker using Inamura capsulorhexis forceps (Duckworth & Kent Ltd., Baldock, UK). Hydrodissection and hydrodelineation were achieved using a balanced salt solution. Phacoemulsification was performed using 2.75 mm-sized phacotips and infusion/aspiration (I/A) cannulas for micro- and small-incision groups, respectively. A clear TIOL (Tecnis ZCT or DIU; Johnson & Johnson Vision, Irvine, CA, USA) was implanted in the capsular bag. The IOL was rotated to the correct axis position according to the axis of total corneal astigmatism. The wound was not sutured. Gatifloxacin 0.3% (Gatiflo, Handok, Chungbuk, Korea) and fluorometholone acetate 0.01% (Ocumetholone, Samil, Seoul, Korea) eye drops were prescribed postoperatively four times a day for four weeks.

### Statistical Analysis

All statistical analyses were performed using a commercial program (SPSS for Windows, version 21.0.1; SPSS Inc., Chicago, IL). The Wilcoxon signed rank test was used to compare pre- and postoperative VA and Corneal endothelial cell density. Comparisons between two groups were performed with Mann–Whitney tests. *P* values < 0.05 were considered statistically significant.

## Results

Table 1 represents the preoperative characteristics of patients in each group. There were no statistically significant differences between the two groups according to age, preoperative best corrected visual acuity (BCVA) (LogMAR), and corneal endothelial cell density (mm^2^) (*p* > 0.05).

**Table 1.**
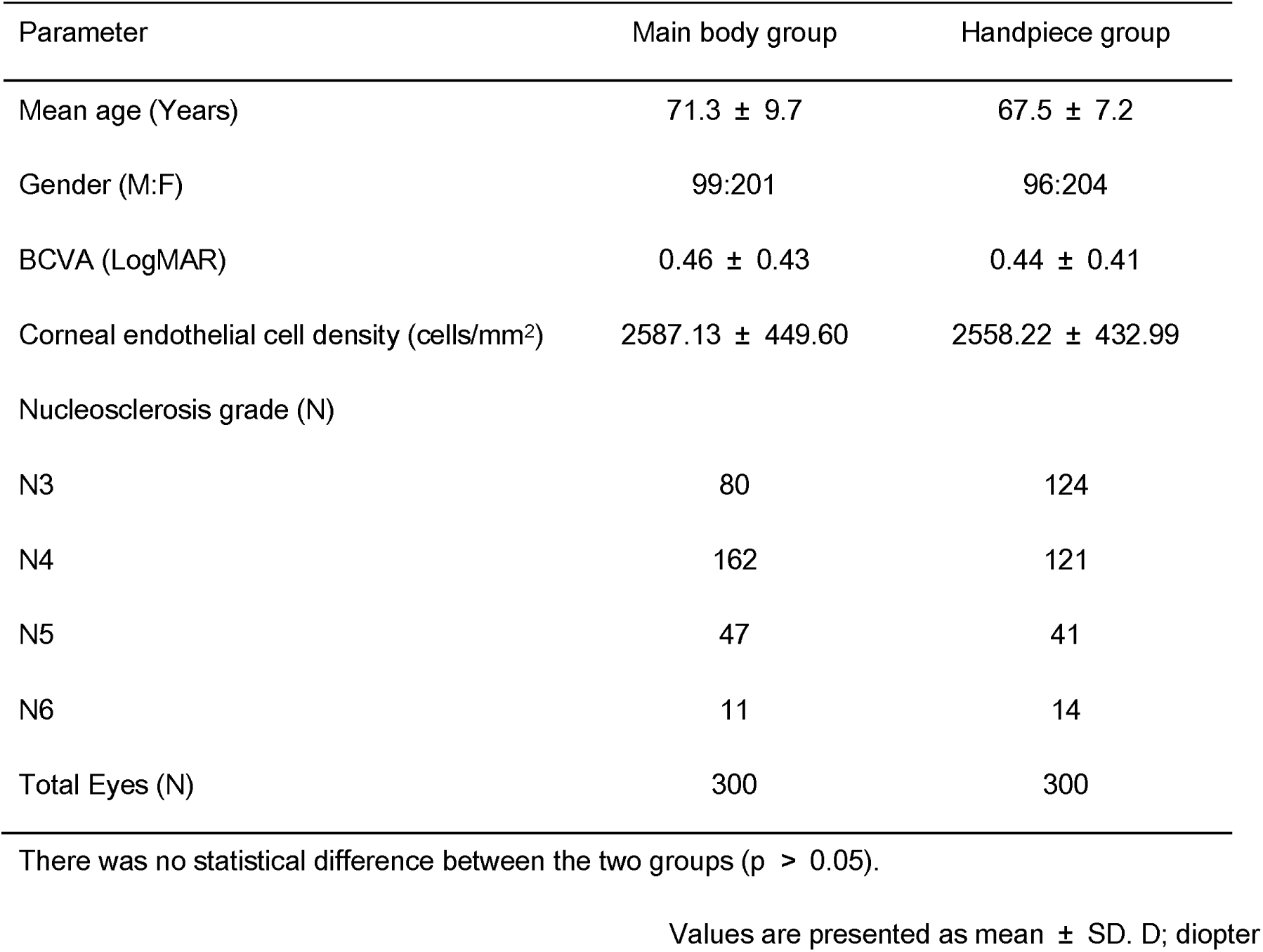
Preoperative data of patients.

| Parameter | Main body group | Handpiece group |
| --- | --- | --- |
| Mean age (Years) | 71.3 ± 9.7 | 67.5 ± 7.2 |
| Gender (M:F) | 99:201 | 96:204 |
| BCVA (LogMAR) | 0.46 ± 0.43 | 0.44 ± 0.41 |
| Corneal endothelial cell density (cells/mm<sup>2</sup>) | 2587.13 ± 449.60 | 2558.22 ± 432.99 |
| Nucleosclerosis grade (N) |  |  |
| N3 | 80 | 124 |
| N4 | 162 | 121 |
| N5 | 47 | 41 |
| N6 | 11 | 14 |
| Total Eyes (N) | 300 | 300 |

There was no statistical difference between the two groups ( $p > 0.05$ ).
Values are presented as mean ± SD. D; diopter

### Postoperative results in handpiece group

Anti-surge (times), phacoemulsification time (seconds), CDE, and BSS use (ml) were significantly increased according to increasing nucleosclerosis grades in the handpiece group, respectively (p < 0.05). Preoperative BCVA (LogMAR) and corneal endothelial cell loss (mm^2^) of nucleosclerosis grades 3 and 4 were worse than grades 1 and 2 (p < 0.05). Postoperative BCVA (LogMAR) of nucleosclerosis grades 4 was worse than grades 1, 2, and 3 (p < 0.05). And, anti-surge (times), phacoemulsification time (seconds), CDE, and BSS use (ml) were significantly increased according to increasing nucleosclerosis grades in the handpiece group, respectively (p < 0.05) (Table 2).

**Table 2.** Postoperative results according to nucleosclerosis grade in handpiece group.

|  | N3 | N4 | N5 | N6 |
| --- | --- | --- | --- | --- |
| Age (years) | 59.23 ± 9.06 | 70.64 ± 8.18 | 73.54 ± 9.62 | 74.71 ± 9.79 |
| Gender (M:F) | 35:89 | 43:78 | 15:26 | 3:11 |
| Anti-surge (times) | 24.38 ± 10.37 | *39.41 ± 15.50 | *44.90 ± 17.05 | *64.50 ± 21.90 |
| Phacoemulsification time (seconds) | 6.38 ± 4.00 | *17.78 ± 7.41 | *38.07 ± 10.28 | *50.92 ± 7.86 |
| CDE | 121.43 ± 32.18 | *264.82 ± 44.25 | *453.93 ± 52.16 | *544.76 ± 58.44 |
| BSS use (ml) | 30.97±11.05 | *48.56±20.06 | *68.27±23.06 | *125.64±45.67 |
| PreOP BCVA (LogMAR) | 0.34 ± 0.30 | 0.42 ± 0.31 | *0.58 ± 0.52 | *1.14 ± 0.72 |
| PostOP BCVA (LogMAR) | 0.07 ± 0.09 | 0.11 ± 0.10 | *0.18 ± 0.13 | *0.24 ± 0.18 |
| Corneal endothelial cell loss (cells/mm<sup>2</sup>) | 213.00 ± 54.29 | 230.64 ± 59.37 | *294.61 ± 50.13 | *311.24 ± 55.39 |
| Eyes (N) | 124 | 121 | 41 | 14 |

CDE = cumulative dissipated energy.
LogMAR = Logarithm of the Minimum Angle of Resolution.
Anti-surge (times), phacoemulsification time (seconds), CDE, and BSS use (ml) of the handpiece group were significantly increased according to increasing nucleosclerosis grades, respectively (p < 0.05).
\*: P<0.05
Values are presented as mean ± SD. D; diopter

We compared the postoperative results according to special situations in handpiece group. Anti-surge (times), phacoemulsification time (seconds), CDE, and BSS use (ml) of the handpiece group were significantly increased in patients with zonule weakness, poor mydriasis, and pseudoexfoliation syndrome compared with those without, respectively (p < 0.05). But there were no statistically significant differences of the anti-surge (times), phacoemulsification time (seconds), CDE, and BSS use (ml) in patients with shallow anterior chamber compared with those without (p > 0.05) (Table 3).

**Table 3.** Postoperative results according to complicated situations in handpiece group.

|  |  | Anti-surge (times) | Phacoemulsification time (seconds) | CDE | BSS use (ml) |
| --- | --- | --- | --- | --- | --- |
| Zonule weakness | Yes (n=44) | *44.43±19.96 | *28.00±15.65 | *336.82 ± 48.29 | *67.05±35.03 |
|  | No (n=256) | 33.52±16.67 | 15.56±13.37 | 186.72 ± 35.36 | 44.23±26.31 |
| Shallow anterior chamber | Yes (n=26) | 35.04±15.50 | 17.77±14.25 | 213.24 ± 35.45 | 46.96±23.18 |
|  | No (n=274) | 35.13±17.80 | 17.35±14.43 | 208.28± 32.25 | 47.64±29.37 |
| Poor mydriasis | Yes (n=25) | *43.92±21.60 | *26.06±17.88 | *312.72± 45.48 | *73.88±45.08 |
|  | No (n=275) | 34.32±17.00 | 16.60±13.80 | 201.28 ± 36.58 | 45.19±25.72 |
| Pseudoexfoliation syndrome | Yes (n=14) | *42.25±24.85 | *26.80±21.33 | *321.61 ± 35.47 | *67.50±46.23 |
|  | No (n=286) | 35.02±17.51 | 17.26±14.28 | 207.12± 31.27 | 47.31±28.57 |

CDE = cumulative dissipated energy
Anti-surge (times), phacoemulsification time (seconds), CDE, and BSS use (ml) of the handpiece group were significantly increased in patients with zonule weakness, poor mydriasis, and pseudoexfoliation syndrome compared with those without, respectively ( $p < 0.05$ ).
\*: $P < 0.05$
Values are presented as mean ± SD. D; diopter

### Comparison between two groups

There were no statistically significant differences between the two groups according to BCVA change (LogMAR) and corneal endothelial cell density (cells/mm^2^) at 3 months after cataract surgery (p > 0.05) (Table 4). But phacoemulsification time (seconds), cumulative dissipated energy (CDE), and BSS use (ml) of the handpiece group (17.39 ± 14.39, 218.72 ± 59.52, and 47.58 ± 28.85, respectively) were significantly lower than those of the main body group (20.44±15.11, 264.84 ± 57.25, and 63.96±42.06, respectively) (p < 0.05) (Figure 1).

**Figure 1.**
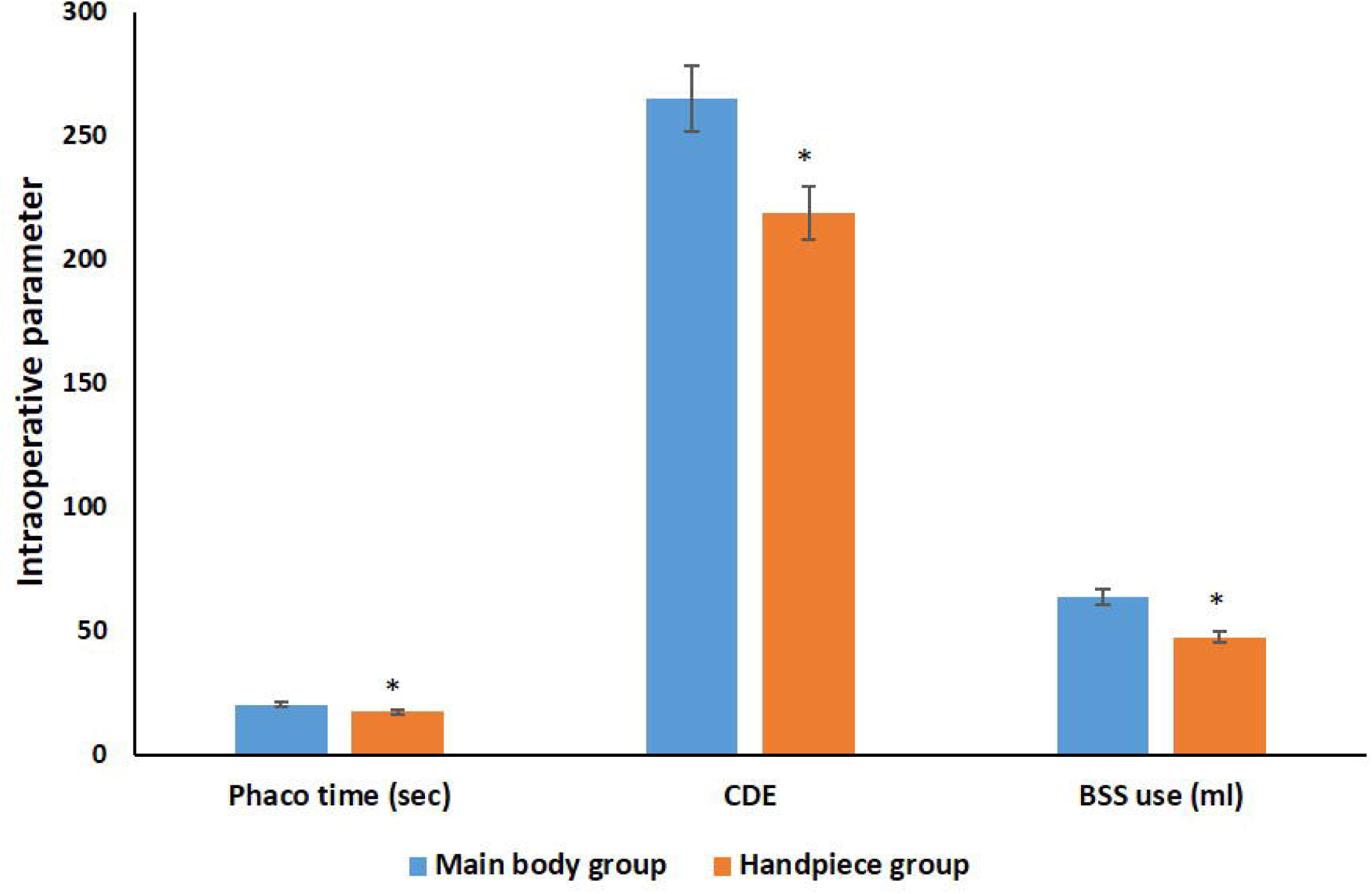
Intraoperative parameters between two groups. Phaco=Phacoemulsification CDE = cumulative dissipated energy BSS = Balanced salt solution Phacoemulsification time (seconds), cumulative dissipated energy (CDE), and BSS use (ml) of the handpiece group were significantly lower than those of the main body group, respectively (p < 0.05). *: P<0.05 Values are presented as mean ± SD.

**Table 4.**
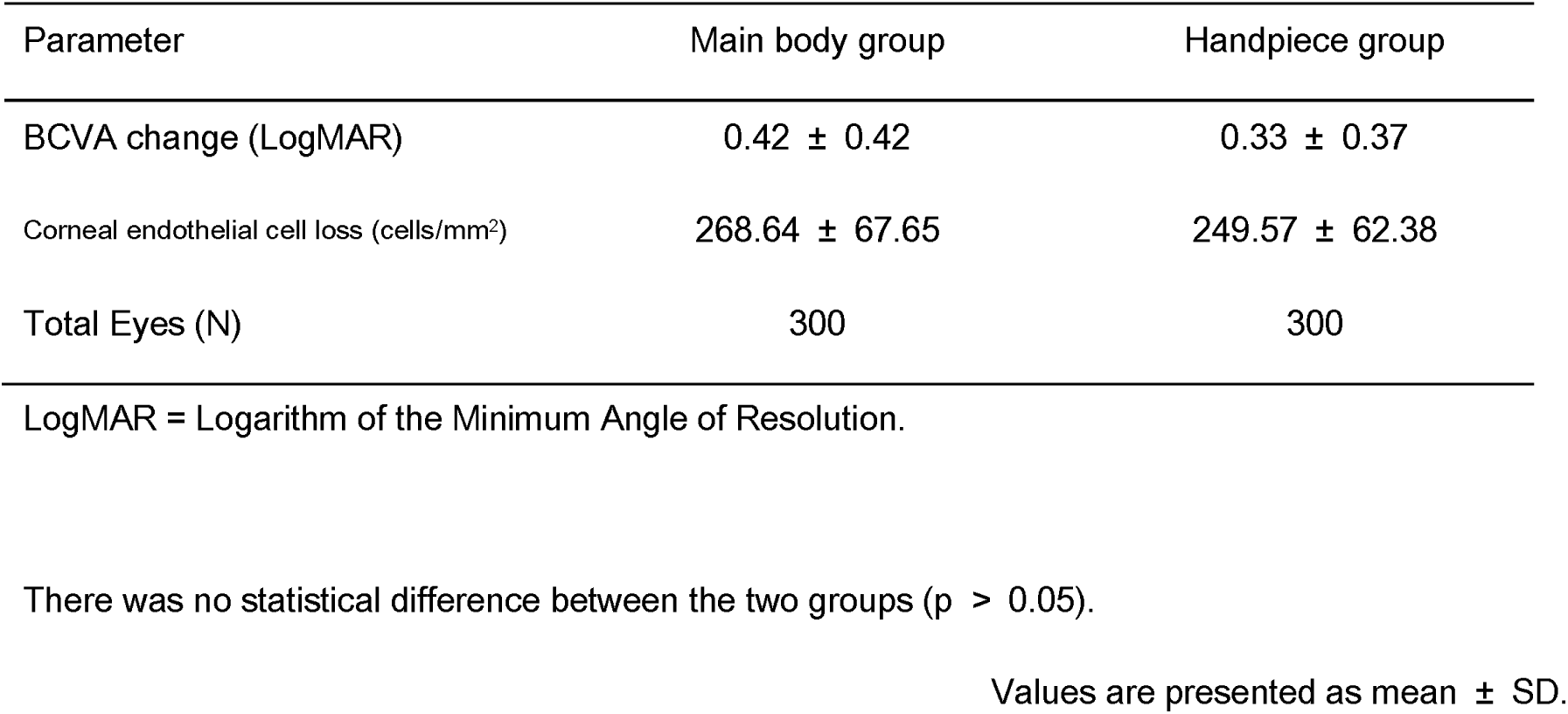
Comparison between main body and handpiece group at 3months after cataract surgery.

| Parameter | Main body group | Handpiece group |
| --- | --- | --- |
| BCVA change (LogMAR) | 0.42 ± 0.42 | 0.33 ± 0.37 |
| Corneal endothelial cell loss (cells/mm<sup>2</sup>) | 268.64 ± 67.65 | 249.57 ± 62.38 |
| Total Eyes (N) | 300 | 300 |

LogMAR = Logarithm of the Minimum Angle of Resolution.
There was no statistical difference between the two groups ( $p > 0.05$ ).
Values are presented as mean ± SD.

We compared the postoperative results between the two groups according to nucleosclerosis grades. There was no statistically significant difference in BCVA change between the two groups according to nucleosclerosis grades (p > 0.05). Phacoemulsification time (seconds), CDE, and corneal endothelial cell loss (cells/mm^2^) of the handpiece group were significantly lower than those of main body group in nucleosclerosis grades 5 and 6 (p < 0.05). BSS uses (ml) of the handpiece group (30.97 ± 11.05, 48.56 ± 20.06, 68.27 ± 23.06, and 125.64 ± 45.67, respectively) were significantly lower than those of the main body group (38.66 ± 18.66, 62.48 ± 37.20, 91.76 ± 37.33, and 153.45 ± 59.24, respectively) in nucleosclerosis grades 3, 4, 5, and 6 (p < 0.05) (Table 5).

**Table 5.** Comparison between main body and handpiece group according to nucleosclerosis grade.

| Nucleo-sclerosis | Group | Phacoemulsification time (seconds) | CDE | BSS use (ml) | BCVA change (LogMAR) | Corneal endothelial cell loss (cells/mm<sup>2</sup>) |
| --- | --- | --- | --- | --- | --- | --- |
| N3 | Main body (n=80) | 8.89 ± 5.02 | 138.21 ± 30.52 | 38.66 ± 18.66 | 0.25 ± 0.22 | 164.34 ± 56.28 |
|  | Handpiece (n=124) | 8.38 ± 4.00 | 121.43 ± 32.18 | *30.97 ± 11.05 | 0.28 ± 0.27 | 213.51 ± 54.29 |
| N4 | Main body (n=162) | 18.33 ± 10.51 | 278.62 ± 63.78 | 62.48 ± 37.20 | 0.37 ± 0.35 | 216.57 ± 55.26 |
|  | Handpiece (n=121) | 17.78 ± 7.41 | 264.82 ± 44.25 | *48.56 ± 20.06 | 0.31 ± 0.28 | 230.64 ± 59.37 |
| N5 | Main body (n=47) | 49.57 ± 11.95 | 498.25 ± 58.12 | 91.76 ± 37.33 | 0.66 ± 0.51 | 427.48 ± 53.87 |
|  | Handpiece (n=41) | *38.07 ± 10.28 | *453.93 ± 52.16 | *68.27 ± 23.06 | 0.39 ± 0.47 | *294.61 ± 50.13 |
| N6 | Main body (n=11) | 63.63 ± 11.08 | 653.54 ± 69.57 | 153.45 ± 59.24 | 1.30 ± 0.43 | 576.86 ± 83.65 |
|  | Handpiece (n=14) | *50.92 ± 7.86 | *544.76 ± 58.44 | *125.64 ± 45.67 | 0.90 ± 0.82 | *311.24 ± 55.39 |

CDE = cumulative dissipated energy
Phacoemulsification time, CDE, and corneal endothelial cell loss of the handpiece group were significantly lower than those of the main body group in nucleosclerosis grades 5 and 6 ( $p < 0.05$ ). BSS uses of the handpiece group were significantly lower than those of the main body in all nucleosclerosis grades ( $p < 0.05$ ).
\*: $P < 0.05$
Values are presented as mean ± SD. D; diopter

## Discussion

Postocclusion surge after occlusion break during phacoemulsification can produce anterior chamber instability. Anterior chamber shallowing and complications such as posterior capsule rupture and vitreous loss can occur due to anterior chamber instability.^9^ Occlusion break surge volumes varied from 17.4 mL to 153 mL, corresponding to 7% and 61%, respectively, of the aqueous volume in the average phakic eye and to 4% and 33% of the aqueous volume in the average aphakic eye.^14^ The magnitude of the postocclusion surge can be reduced with low-compliance tubing, aspiration bypass systems (venting), modified phaco needles, and low infusion pressure.^8 15 16^ So, surgeons can safely increase the vacuum levels used during phacoemulsification.^5 9^

The active-fluidics system (AFS) can allow the pre-setting of an IOP value and maintain the target IOP through regulating the irrigation pressure compared with the conventional gravity-based fluidic system (GFS).^3 17^ So, AFS can improve the efficiency, effects, safety, and patients’ subjective perceptions of phacoemulsification compared with GFS.^18^ Some studies have reported that the AFS conserved CDE with a variation of 19%–40%.^17 19 20^ But other studies have reported that CDE might be related to the surgical techniques, incorporating the severity of the patients’ condition.^21 22^

Active Fluidics was equipped with the Active Sentry (Alcon Laboratories, Inc.), which contains an integrated pressure sensor in the fluidic management system to control the postocclusion surge with the venting system.^10^ The Active Sentry system was reported to be useful for maintaining the anterior chamber depth even during occlusion breaks compared with other systems.^1 4 23^ And the Centurion Vision System has been upgraded, where the IOP within the anterior chamber is sensed at the level of the handpiece instead of the fluidic management system.^24^ Kim JY et al. reported that active surge mitigation (ASM) actuation increases with age, poor visual acuity before surgery, short axial length, and prolonged ultrasound usage time.^13^ They expected that the Active Sentry handpiece would function more effectively in more severe and high-risk cataract surgery.^13^ Sabur H. et al. reported that ASM actuations were more engaged in the eyes with pseudoexfoliation and small pupils.^25^ In our results, Anti-surge (times), phacoemulsification time (seconds), CDE, and BSS use (ml) of the handpiece group were significantly increased in patients with zonule weakness and poor mydriasis compared with patients without, respectively (p < 0.05) (Table 3). And, anti-surge (times), phacoemulsification time (seconds), CDE, and BSS use (ml) were significantly increased according to increasing nucleosclerosis grades in the handpiece group, respectively (p < 0.05) (Table 2).

There were several studies to compare the Active Sentry handpiece and the traditional Ozil handpiece.^10 12 13 24^ Kim JY et al. reported that there was no clear advantage of the Active Sentry handpiece compared to the Centurion Ozil handpiece.^13^ Cyril et al. reported that surgeons were more comfortable using the Ozil handpiece during entry into the anterior chamber and emulsification of hard nuclei (48.5% vs. 28.6%). The mean cumulative dissipated energy for soft and hard cataracts was 5.6 and 4.8 and 9.3 and 9.4 for the Ozil and Active Sentry groups, respectively.^10^ But in our results, the CDE and corneal endothelial cell loss (mm^2^) of the handpiece group were significantly lower than those of the main body group in nucleosclerosis grades 5 and 6 (p < 0.05) (Table 5). Sabur H. et al. also reported that CDE and torsional amplitude measured in the Active Sentry handpiece group (8.8 ±3.9, 51.2 ±13.3, respectively) were significantly lower than those of the Ozil handpiece group (10.4±4.2, 65.2±9.3, respectively).^25^

Vasavada et al. reported that the rise of IOP to baseline after the occlusion break event was faster and the mean percentage reduction of IOP from maximum during nuclear fragment removal was lower when using the Centurion Vision system with the Active Sentry upgrade compared with the traditional handpiece.^24^ In our results, the phacoemulsification time (seconds) of the handpiece group was significantly lower than that of the main body group in nucleosclerosis grades 5 and 6 (p < 0.05) (Table 5).

Jirásková N. et al. reported that the mean CDE and the phacoemulsification time were significantly statistically decreased using Active Sentry versus Centurion Ozil handpieces.^12^ In this result, phacoemulsification time (seconds), cumulative dissipated energy (CDE), and BSS use (ml) of the handpiece group were significantly lower than those of the main body group, respectively (p < 0.05)(Figure 1). Jirásková N. et al. reported that the difference in estimated consumption of balanced salt solutions was not statistically significant.^12^ But this was not analyzed according to nucleosclerosis grade. In our results, BSS uses (ml) of the handpiece group (30.97 ± 11.05, 48.56 ± 20.06, 68.27 ± 23.06, and 125.64 ± 45.67) were significantly lower than those of the main body group (38.66 ± 18.66, 62.48 ± 37.20, 91.76 ± 37.33, and 153.45 ± 59.24, respectively) in nucleosclerosis grades 3, 4, 5, and 6 (p < 0.05) (Table 5).

To the best of our knowledge, this is the first study about the comparison of the efficacy of the Centrion handpiece anti-surge system and main body according to the grade of nucleosclerosis and complicated situations such as pseudoexfoliation syndrome, zonule weakness, and poor mydriasis In our study, there was a limitation that a multicenter clinical trial with a larger sample size and a longer follow-up period was needed to observe the long-term efficacy of Centrion handpiece anti-surge system and main body.

In conclusion, Anti-surge times increased according to increasing nucleosclerosis grades and in patients with zonule weakness, poor mydriasis, and pseudoexfoliation syndrome. Because of the fast-reacting anti-surge, phacoemulsification time, CDE, and BSS use (ml) were saved in the handpiece anti-surge system rather than in the main body. Phacoemulsification time (seconds), CDE, and CEC loss (cells/mm^2^) of the handpiece group were also significantly lower than those of the main body group in patients with hard nuclei. Therefore, the anti-surge system in the Centrion handpiece helps the surgeon perform safer cataract surgery in higher nucleosclerosis grade, zonule weakness, poor mydriasis, and pseudoexfoliation syndrome. The ability to sense IOP at the level of the handpiece with the Active Sentry upgrade allows faster mitigation of surge response.^24^

## Data Availability

All data produced in the present study are available upon reasonable request to the authors

## Notes

This work was supported by the National Research Foundation of Korea (NRF) grant funded by the Korea government (MSIT) (No. 2022R1F1A1069218) and Alcon through an Investigator Initiated Trial (IIT No. 73949933).

The authors do not have any conflicts of interest or proprietary interests in the contents of this manuscript.

### Competing Interest Statement

The authors have declared no competing interest.

### Author Declarations

The Institutional Review Board (IRB)/Ethics Committee of Bucheon St. Mary Hospital approved this study protocol.

